# Development and Validation of an Obstetric Early Warning System model for use in low resource settings

**DOI:** 10.1101/2020.07.31.20165209

**Authors:** Aminu Umar, Alexander Manu, Matthews Mathai, Charles Ameh

## Abstract

**Background:** The use of obstetric early-warning-systems (EWS) has been recommended to improve timely recognition, management and early referral of women who have or are developing a critical illness. Development of such prediction models should involve a statistical combination of predictor clinical observations into a multivariable model which should be validated. No obstetric EWS has been developed and validated for low resource settings. We report on the development and validation of a simple prediction model for obstetric morbidity and mortality in resource-limited settings.

**Methods:** We performed a multivariate logistic regression analysis using a retrospective case-control analysis of secondary data with clinical indices predictive of severe maternal outcome (SMO). Cases for design and validation were randomly selected (n=500) from 4360 women diagnosed with SMO in 42 Nigerian tertiary-hospitals between June 2012 and mid-August 2013. Controls were 1000 obstetric admissions without SMO diagnosis. We used clinical observations collected within 24 hours of SMO occurrence for cases, and normal births for controls. We created a combined dataset with two controls per case, split randomly into development (n=600) and validation (n=900) datasets. We assessed the model’s validity using sensitivity and specificity measures and its overall performance in predicting SMO using receiver operator characteristic (ROC) curves. We then fitted the final developmental model on the validation dataset and assessed its performance. Using the reference range proposed in the United Kingdom Confidential-Enquiry-into-Maternal-and-Child-Health 2007-report, we converted the model into a simple score-based obstetric EWS algorithm.

**Results:** The final developmental model comprised abnormal systolic blood pressure-(SBP>140mm Hg or <90mmHg), high diastolic blood pressure-(DBP>90mmHg), respiratory rate-(RR>40/min), temperature-(>38°C), pulse rate-(PR>120/min), caesarean-birth, and the number of previous caesarean-births. The model was 86 % (95% CI 81-90) sensitive and 92%-(95% CI 89-94) specific in predicting SMO with area under ROC of 92% (95% CI 90% – 95%). All parameters were significant in the validation model except DBP. The model maintained good discriminatory power in the validation (n=900) dataset (AUC 92, 95% CI 88-94%) and had good screening characteristics. Low urine output (300mls/24hours) and conscious level (prolonged unconsciousness-GCS<8/15) were strong predictors of SMO in the univariate analysis.

**Conclusion:** We developed and validated statistical models that performed well in predicting SMO using data from a low resource settings. Based on these, we proposed a simple score based obstetric EWS algorithm with RR, temperature, systolic BP, pulse rate, consciousness level, urinary output and mode of birth that has a potential for clinical use in low-resource settings.

## Background

The World Health Organization (WHO) estimated that 303,000 maternal deaths occurred globally at the end of the Millennium Development Goals (MDGs) in 2015. Over 99% of these deaths occurred in low and middle-income countries (LMICs), most of which made insufficient progress towards achieving the MDG maternal health targets ^1^. It is also estimated that there are 27 million episodes of direct obstetric complications annually which contribute to long-term pregnancy and childbirth complications ^2^. Following increasing access to facility-based births, partly because of Universal Health Coverage policies under the Sustainable Development Goals since 2016, opportunities to ensure good quality facility care are critical, if the new ambitious global and national Maternal Mortality Ratio (MMR) targets are to be achieved ^3^.

The increased burden of adverse outcomes, especially in LMICs is believed to be due primarily to delays in the recognition of pregnancy complications ^4,5^. Early warning systems (EWSs) are clinical diagnostic prediction models that involve serial clinical observations (“track”) with criteria (“trigger”) to identify patients at risk of complications ^6^. A 2018 systematic review of EWS used in obstetrics found that they are effective in predicting adverse obstetric outcomes and reducing obstetric morbidity^7^

The United Kingdom Confidential Enquiry into Maternal and Child Health (CEMACH) (2003-2005 report) recommended “the use of obstetric EWS to improve timely recognition, treatment and referral of women who have or are developing a critical illness” ^8^. Most of the available obstetric EWS versions used subsequently were designed based on clinical consensus rather than the application of recommended prediction model development methodology that should include statistical analysis of outcome measures ^9–17^.

Model development involves a statistical combination of predictor clinical observations into a multivariable model. The Transparent Reporting of a multivariable prediction model for Individual Prognosis Or Diagnosis (TRIPOD) statement recommends that new prediction models are tested on data used in its development (internal validation) and data from a different population (external validation) ^10^.

In 2013, the first statistically derived, obstetric EWS was developed in the UK. It was internally validated using clinical observations (physiological variables) collected from 4400 women during their first 24 hours of critical care admission ^18^. The EWS developed showed a good predictive ability to discriminate survivors from non-survivors in the derivation dataset, as well as on an external dataset ^19^. However, since the database used in the development, internal ^18^ and external validation ^19^ of the EWS were only for women admitted to critical care, the EWS may not be suitable for obstetric populations without obvious need for critical care and in a non-UK setting.

No obstetric EWS has been developed and validated for a low resource setting. However, 14 of the 16 obstetric EWS identified in a recent systematic review, had five clinical observations of physiologic variables (pulse rate, systolic and diastolic blood pressure, respiratory rate, temperature and consciousness level) that can be easily collected even in low resource settings.^7^

We report on the development and validation of a simple obstetric prediction model and EWS algorithm for use in resource-limited settings.

## Methods

### Study design

The study was a retrospective case-control analysis of secondary data on admissions to inpatient obstetric wards in Nigerian tertiary hospitals. Cases based on standardised definitions were derived from the Nigerian near-miss study, the largest prospective investigation of maternal deaths and near-misses in Africa.^20^. Controls were women who were admitted for obstetric care at the same time as the cases.

### Study Population and Data Sources

The study protocol and findings of the Nigerian Near-miss study have been published elsewhere ^20,21^. All women admitted to 42 Nigerian tertiary hospitals for birth or within 42 days of birth or termination of pregnancy between June 2012 and August 2013 were eligible for enrolment into the near-miss study. The study reported severe maternal outcome (SMO) cases based on the WHO near-miss criteria (organ dysfunction, clinical and management-based) ^20,22^.

For this analysis, we obtained data on 4360 women who had SMO during the 14-month surveillance, including 998 who died and 3362 who had a near-miss ^22^. Overall, 2401 SMO cases occurred in 11 tertiary care hospitals across the 3 northern sub-regions of Nigeria. Cases (n=500) were randomly selected from these, and split into the design (n = 200) and validation (n = 300) SMO cases dataset (Figure 1). Three of the 11 tertiary care hospitals were coveniently selected across the three sub-regions (one tertiary hospital in each northern sub-region) for control data collection. One thousand women who were admitted for birth, between June 2012 and August 2013, when the near-miss study was conducted and discharged with no SMO were recruited as the controls.

**Figure 1:**
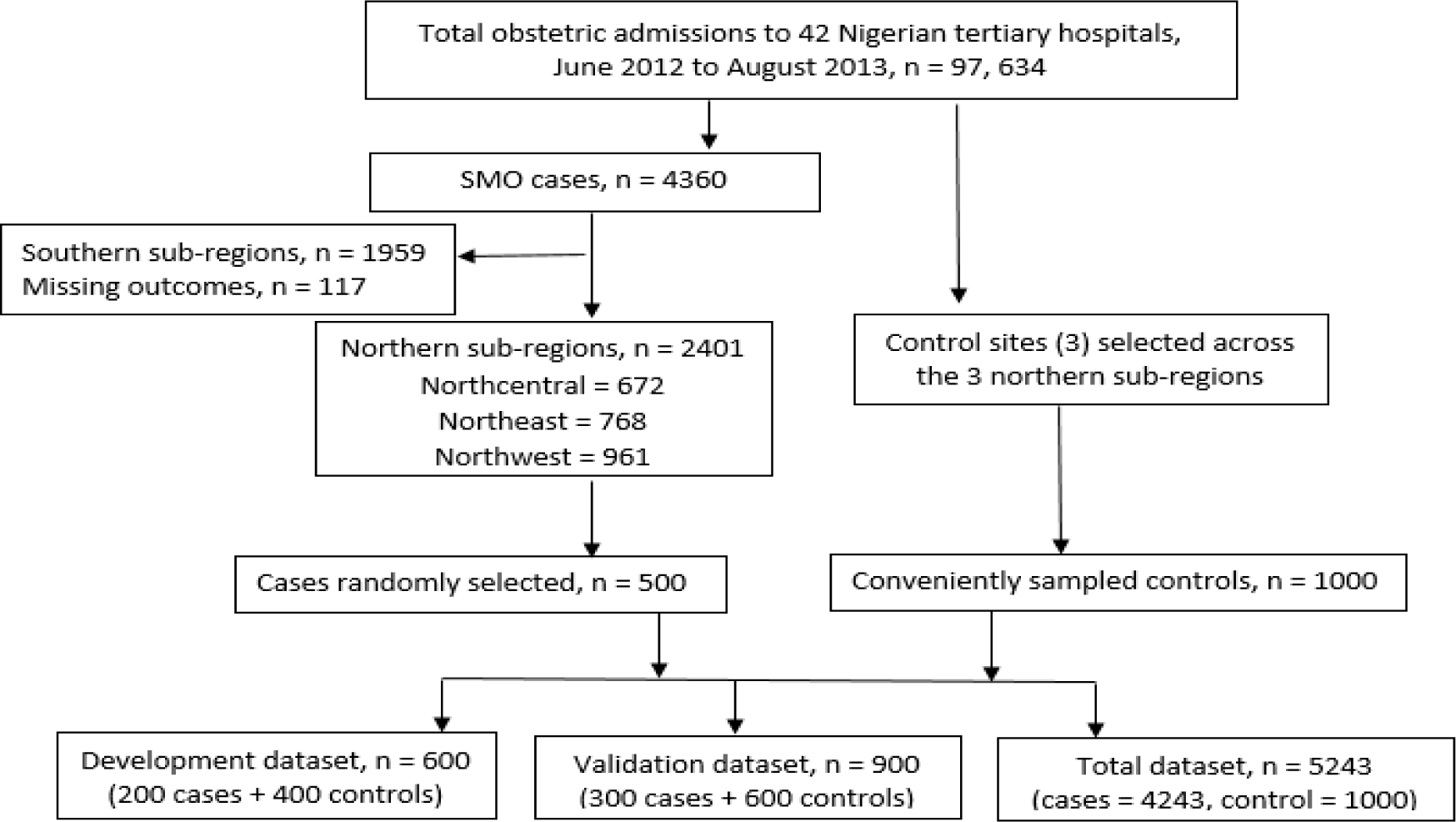
Creation of the development and validation data sets.

The sample size estimate was based on a baseline SMO prevalence of 4·8% from the near-miss case dataset. At 5% level of significance, the analysis could detect an absolute difference in SMO prevalence of 6% and 5% at 90% and 80% powers, respectively.

All data were collected within 24 hours of the occurrence of SMO (for cases) (Oladapo et al, 2013) or 24 hours of birth or end of pregnancy (collected retrospectively for controls). Figure 1 illustrates allocation into, and composition of, the development and validation data sets.

### Data abstraction

Individual-level data on all study variables were abstracted from the Near-miss study dataset (n=4360) and from the case notes of 1000 controls. These included demographic characteristics (age, weight, height), obstetric variables (parity, the number of antenatal clinic visits, gestational age at the time of admission or birth, mode of birth, interval from last pregnancy, the number of previous caesarean section), diagnosis, length of stay in hospital, and the last haematocrit measured before occurrence of outcome. Abnormal clinical indices were extracted from the cases dataset based on their definitions codebook including high (>140 mm Hg) and low (<90 mm Hg) systolic blood pressure, high (>90 mm Hg) and low (<60 mm Hg) diastolic blood pressure, high (>38°C) and low (<35°C) temperature, marked tachycardia (PR>120/min) and bradycardia (PR<60/min), hypoxaemia (SpO_2_<90%), severe tachypnoea (RR>40/min) and bradypnea (RR<6/min), severe oliguria (urinary output<300 mL in 24 hours), and coma (Glasgow Coma Score<8/15). Among controls, data were collected as continuous variables and classified based on the cut-off values in the cases dataset codebook.

### Outcome

The outcome measure for this analysis was SMO, computed as the sum of maternal deaths and near-misses and transformed into a binary variable: occurred or not. Maternal death was defined according to the International Classification of Diseases (ICD-10) ^23^. Women were identified as maternal near-miss if they met any of the three near-miss criteria (clinical criteria related to specific disease entities, intervention-based criteria and organ dysfunction-based criteria) ^22^.

### Statistical methods

Characteristics of the study population were summarised by means and standard deviations for continuous and percentages for categorical variables. Continuous variables were compared between cases and controls with independent sample t-tests or Mann Whitney U tests, depending on whether or not variables were normally distributed. Normality was assessed visually through distribution plots and with the Shapiro-Wilk test. Data for 1500 (cases = 500 and controls, n = 1000) study participants were randomly allocated into the development dataset (n=600) or validation data set (n=900) with two controls per case.

### Model building

Univariable logistic regression models were fitted to assess the association between each predictor variable and outcome (SMO). A stricter inclusion criterion was applied and therefore variables were only selected from the univariable models for inclusion into the multivariable model if the model had a p-value<0·05. This yielded 22 potential variables for inclusion.

Multivariable logistic regression models were fitted using a backward stepwise approach and factors were removed from the model, one at a time based on the highest p-value>0·05 and their likelihood ratio. When the final model was achieved, a sensitivity test was performed by including each of the eliminated variables, in turn, into the final model to assess their significance. None of these was found significant in the final model. Variables were tested for collinearity, by a simple check of the correlation coefficients, and were dropped to improve parsimony of the model.

### Model performance

Performance of the obstetric EWS clinical prediction model from the development data set was tested on the validation data set ^24^. First, overall validity (sensitivity, specificity, negative predictive value-NPV and positive predictive value-PPV), as well as area under the curve (AUC) for Receiver Operating Characteristics (ROC) curves were assessed for the final model. Discrimination was assessed using the *c* statistic or AUC, an estimate of the probability of assigning higher risk to those who suffered SMO compared to those who did not. The final model was then applied to the validation dataset, and performance was similarly assessed. Finally, we estimated validity of the selected EWS predictors by applying the final (developmental) model to the entire (n=5243 after excluding 117 missing data) dataset of cases and controls, bearing in mind that the ratio of cases to controls had been reversed – over 4 cases per control. All statistical analyses were performed using Statistical Package for the Social Sciences (SPSS) software version 25 and Stata version 15·1. We constructed 95% confidence intervals (CI) for all performance characteristics.

### Obstetric EWS algorithm

Our statistically derived diagnostic prediction model was modified based on the clinical importance of rejected variables. Given that the cases data set provided categorised (binary) clinical variables, it was not feasible to validate reference ranges for the different model parameters. Hence, the final (validation) model was converted into simple score-based obstetric EWS algorithm using the reference range proposed in the MEOWS chart recommended in the 2007 CEMACH report ^8^ (Additional file: Appendix 1).

### Patient and Public Involvement statement

Patients were not involved in the development of the research question or design of this study. Secondary patient monitoring data was used for this analysis. Implications of this study will be disseminated through patient groups and blogs in the study setting.

We present our findings according to the Transparent Reporting of a multivariable prediction model for Individual Prognosis Or Diagnosis statement^10^.

## Results

From the 97, 634 women admitted for pregnancy, childbirth or puerperal complications during the 14 months of the Nigerian near-miss study, 4360 developed SMO and their data were included in the cases data set. Of these, 998 women died and 3362 suffered maternal near-miss. A total of 117 (2·7%) women with SMO had missing data, with no records of the type of SMO they experienced. These women were excluded from the analyses as the missingness was assumed to be at random. Lack of data on other characteristics of these participants did not allow for assessment of bias in dropping them from the analysis. None of the control participants had missing parameters.

Characteristics of SMO cases and controls are given in Table 1. Those who experienced SMO tended to be older and more likely to be obese. They also stayed longer in hospital, with the mean number of days of admission four times greater than controls and were more likely to be anaemic. There was no significant difference in terms of parity, but SMO cases had more preterm births and had more antenatal visits than the controls (Table 1).

**Table 1:**
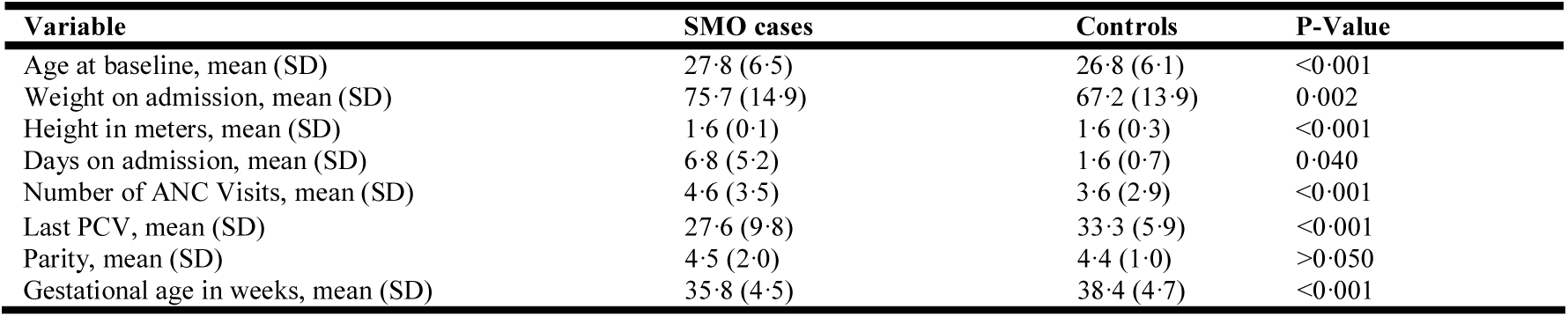
Characteristics of women with severe maternal outcome (n=4243) compared with controls who were discharged without SMO diagnosis (n=1000)

Overall, women who had abnormal clinical observation measurements (either lower or higher than normal), based on the pre-defined cut-off points deciphered from the cases dataset, were more likely to develop SMO than controls (Table 2). Initially, bivariate analysis was performed considering all 22 potential variables for entry in the model. Of these, a total of 15 had significant p-values <0·05 (Table 2) and so these were considered for potential inclusion in the multiple regression model.

**Table 2:** Statistically significant clinical variables from univariate analysis (dependent variable; SMO binary outcome variable) in the model design dataset (n=600)

| Parameters | Cases<br>N=200 | Controls<br>N=400 | Chi-square<br>significance |
| --- | --- | --- | --- |
| High systolic blood pressure (>140), *number (%)* | 75 (68.8) | 34 (31.2) | <0.001 |
| Low systolic blood pressure (<90), *number (%)* | 58 (61.7) | 36 (38.3) | <0.001 |
| High diastolic blood pressure (>90), *number (%)* | 75 (64.7) | 41 (35.3) | <0.001 |
| Low diastolic blood pressure (<60), *number (%)* | 110 (65.5) | 58 (34.5) | <0.001 |
| Severe tachypnoea (RR>40), *number (%)* | 26 (92.9) | 2 (7.1) | <0.001 |
| Severe bradypnea (RR<6), *number (%)* | 5 (100) | 0 (0) | <0.001 |
| Fever (Temp>38), *number (%)* | 17 (94.4) | 1 (5.6) | <0.001 |
| Marked tachycardia (PR>120), *number (%)* | 58 (71.6) | 23 (28.4) | <0.001 |
| Hypoxaemia (SP02<90%), *number (%)* | 14 (82.4) | 3 (17.6) | <0.001 |
| Caesarean delivery in present admission, *number (%)* | 51 (94.4) | 3 (5.6) | 0.001 |
| Low urinary output (300ml/24hours), *number (%)* | 12 (100) | 0 (0) | <0.001 |
| Prolonged unconsciousness (GCS<8/15), *number (%)* | 14 (100) | 0 (0) | <0.001 |
| Blood transfusion in present admission, *number (%)* | 40 (67.8) | 19 (32.8) | 0.038 |
| Last haematocrit level, *mean (SD)* | 27.6 (9.8) | 33.3 (5.9) | <0.001 |
| Days of admission, *mean (SD)* | 6.8 (5.2) | 1.6 (0.7) | 0.040 |

The significant variables were entered into a multiple logistic regression model. Both backward and forward stepwise selection methods were used to build the final parsimonious model, with the standard 5% significance level for entry and removal. Both techniques produced the same final (developmental) model, in five steps, with 8 parameters (high systolic blood pressure (>140 mm Hg), low systolic blood pressure (<90 mm Hg), high diastolic blood pressure (>90 mm Hg), severe tachypn0ea (RR>40/min), fever (temperature >38° C), marked tachycardia (PR>120/min), mode of birth (caesarean or vaginal birth), and the number of previous caesarean births).

### Risk of severe maternal outcome

**Table 3** gives odds ratios of developing SMO for the variables in the SMO models. From the developmental model, risk of developing SMO was five times greater among women with high systolic blood pressure (>140 mm Hg) and tachycardia (PR>120/min), while low systolic BP (<90 mm Hg) increased SMO risk four times. Caesarean birth during index admission was found to increase risk of SMO significantly (odds ratio 6; 95% CI 2.8-12.5), but the number of previous caesarean sections did not increase risk. Most importantly, the variables with the highest risk of SMO in the developmental model were fever (OR 116.5; 95% CI 13.0-147.4) and tachypnoea, (OR 25.2; 95% CI 4.2-51.6).

**Table 3:** Odds ratios for SMO of the significant predictor variables.

| **Developmental model (n=600; 200 cases versus 400 controls)** |  |  |  |  |  |
| --- | --- | --- | --- | --- | --- |
| **Parameters** | **Odd ratio** | **S. E** | **Significance (p)** | **95% CI** |  |
|  |  |  |  | **Lower** | **Upper** |
| sBP>140 mmHg | 5.26 | 0.49 | 0.001 | 2.03 | 13.63 |
| sBP<90 mmHg | 3.73 | 0.42 | 0.002 | 1.65 | 8.41 |
| dBp>90 mmHg | 2.78 | 0.48 | 0.035 | 1.08 | 7.16 |
| RR>40 cycles/min. | 25.20 | 0.92 | <0.001 | 4.19 | 51.60 |
| Temp>38°C | 116.51 | 1.12 | <0.001 | 12.96 | 147.42 |
| PR>120/min | 4.62 | 0.43 | <0.001 | 2.00 | 10.66 |
| CS (Yes vs No) | 5.91 | 0.38 | <0.001 | 2.79 | 12.53 |
| Number of CS | 0.96 | 0.01 | <0.001 | 0.95 | 0.97 |
| **Validation model (n=900); 300 cases versus 600 controls)** |  |  |  |  |  |
| **Parameters** | **Odd ratio** | **S. E** | **Significance (p)** | **95% CI** |  |
|  |  |  |  | **Lower** | **Upper** |
| sBP >140 mmHg | 8.61 | 0.56 | <0.001 | 4.32 | 32.82 |
| sBP < 90 mmHg | 5.42 | 0.33 | 0.002 | 2.66 | 7.35 |
| dBp > 90 mmHg | 2.80 | 0.45 | 0.119 | 0.65 | 8.70 |
| Temp>38°C | 123.21 | 2.95 | 0.002 | 62.83 | 136.95 |
| RR>40 cycles/min | 18.21 | 2.40 | <0.001 | 3.43 | 24.31 |
| CS (yes vs. no) | 5.85 | 1.42 | <0.001 | 3.59 | 11.19 |
| Number of CS | 0.97 | 0.06 | <0.001 | 0.95 | 0.99 |
| PR>120/min | 4.84 | 0.40 | <0.001 | 1.69 | 9.08 |
| **Performance checking model with total data set (n=5243)** |  |  |  |  |  |
| **Parameters** | **Odd ratio** | **S. E** | **Significance (p)** | **95% CI** |  |
|  |  |  |  | **Lower** | **Upper** |
| sBP >140 mmHg | 15.50 | 0.31 | <0.001 | 8.38 | 28.69 |
| sBP < 90 mmHg | 2.09 | 0.20 | <0.001 | 1.42 | 3.08 |
| dBp > 90 mmHg | 0.89 | 0.30 | 0.706 | 0.50 | 1.61 |
| RR > 40 cycles/min | 23.19 | 0.47 | <0.001 | 9.18 | 58.57 |
| Temp > 38°C | 55.90 | 0.46 | <0.001 | 22.64 | 138.02 |
| CS (yes vs no) | 6.38 | 0.16 | <0.001 | 4.71 | 8.65 |
| Number of CS | 0.98 | 0.00 | <0.001 | 0.98 | 0.98 |
| PR > 120 | 6.27 | 0.21 | <0.001 | 4.14 | 9.50 |

\*Absolute risk estimated by rounding the odds ratios to one significant figure

Although significant in the developmental model, high diastolic pressure (>90mmHg) was not as much of a risk factor as systolic BP, and thus ceased to have a significant effect on SMO risk in the validation model and the performance checking model with the whole data (Table 3). When interactions were considered, diastolic blood pressure was found to be strongly collinear (correlation coefficient, r = 0·95) with systolic blood pressure. When applied in the validation data set (n=900) and the whole dataset (n=5243, for sensitivity analysis), all other variables in the developmental model produced a consistent effect in the same direction with variations in effect sizes (Table 3). Similarly, the clinical variables with the highest risk of SMO in the two models were fever and tachypnoea, with odds ratios of 123.2 (95% CI, 62.8-137.0) and 18.2 (95% CI, 3.4-24.3) in the validation model, and 55.9 (95% CI, 22.6-138.0) and 23.2 (95% CI, 9.2-58.6) in the performance checking model with the whole data (Table 3).

### Predictive accuracy for SMO

The developmental model explained 66% of the variability in SMO with AUROC of 92% (Table 4, Additional file: Appendix 3). Given that the cases data set provides already categorised (binary) clinical variables, the diagnostic accuracy of the models was assessed based on the number of parameters required to predict SMO with the best screening properties. Using the presence of five or more triggers as the cut-off point to define SMO, the model predicted SMO with a sensitivity of 86%, a specificity of 92%, positive and negative predictive values of 84% and 93% (Table 4). The validation model produced very similar screening characteristic and discriminatory ability (AUROC 92%) (Table 4, Additional file: Appendix 4). Not surprisingly, however, the sensitivity model with all cases and controls produced the highest positive predictive value (94%), but a significantly reduced negative predictive value (61%). This is expected given the high prevalence of the outcome measure in the data set (case: control = 4:1). The model explained 58% of the variability in SMO with 92% discriminatory ability (AUROC 82%) (Table 4, Additional file: Appendix 5).

**Table 4:**
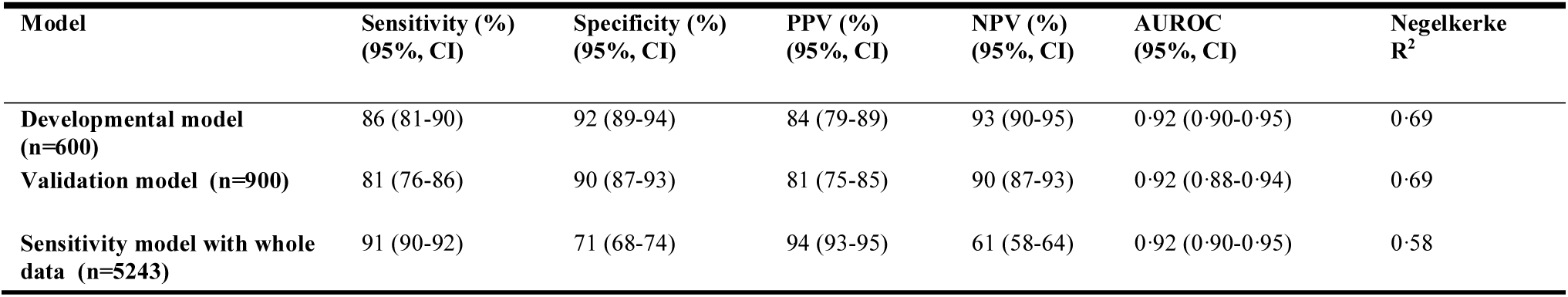
Predictive accuracy of the SMO models (best performing cut-off 0.6)

### Proposed EWS algorithm

Measurements of temperature, pulse rate, systolic blood pressure, respiratory rate and mode of birth in postpartum women (caesarean birth versus vaginal birth), constitute the primary early warning parameters from the three statistical models. Diastolic blood pressure was dropped as it was strongly collinear with systolic blood pressure (r = 0·95), and the latter was more clinically relevant and significant in all statistical models (Table 3). Consciousness level and low urinary output (anuria) were dropped in the statistical models due to the perfect prediction of outcome, not statistical significance; this implied that none of the controls (n=1000) suffered prolong unconsciousness (GCS<8/15) or experienced low urinary output. Although the two variables were strongly significant predictors of SMO at univariate level (Table 2), they were dropped in the statistical models. Therefore, both variables were forced into the proposed obstetric EWS algorithm, adopting the AVPU (alert, responds to voice or pain and unresponsive) for consciousness level from the MEOWS chart recommended in the 2003-2005 CEMACH report (Additional file: Appendices 1 and 2). Defining trigger as a single markedly abnormal observation (red trigger) or the combination of two simultaneously mildly abnormal observations (two yellow triggers), the corresponding values from the CEMACH MEOWS chart (Additional file: Appendix 1) were converted into scores of 0 (normal observation), 1 (yellow trigger) and 2 (red trigger) in the proposed algorithm (Table 5). Mode of birth was scored as 0 and 1 for vaginal and caesarean births.

**Table 5:** Scoring guideline for the proposed obstetric EWS algorithm

| Parameters | 2 | 1 | 0 | 1 | 2 |
| --- | --- | --- | --- | --- | --- |
| **Temperature** | <35 | 35- <36 | 36 - <38 |  | >38 |
| **Pulse rate** | <40 | 40 - <50 | 50 - <100 | 100-120 | >120 |
| **Respiratory rate** | 0 – 10 |  | 11-20 | 21-30 | >30 |
| **Systolic blood pressure** | <90 | 90-<100 | 100 - <150 | 150 – 160 | >160 |
| **Urine volume (ml/hour)** | <20 | 20 – 30 | >30 |  |  |
| **Mode of birth** | CS |  | Vaginal birth |  | CS |
| **Consciousness level**<br>**(AVPU)** | Response to pain/<br>unresponsive | Response to<br>voice | Alert | Response to<br>voice | Response to pain/<br>unresponsive |

## Discussion

This study, to our knowledge, reports for the first time the development and validation of an obstetric diagnostic prediction model for a general obstetric population in a low resource setting using recommended methodology^10^. The final developmental model comprised abnormal systolic blood pressure (SBP>140mm Hg or <90mmHg), high diastolic blood pressure (>90mmHg), respiratory rate (RR>40/min), temperature (>38oC), pulse rate (PR>120/min), caesarean birth, and the number of previous caesarean births. The model was 86% (95% CI 81-90) sensitive and 92% (95% CI 89-94) specific in predicting SMO with AUROC of 92% (95% CI 90 – 95%). We proposed a score-based obstetric EWS algorithm with seven clinical parameters (RR, temperature, systolic BP, pulse rate, consciousness level, urinary output and mode of birth) that has a potential for clinical use in low-resource settings.

A previous study in the United Kingdom using a similar methodology produced an obstetric EWS with similar discriminatory properties (AUROC) 96% (95% CI 92-99%).^18^ Although that study by Carle et al. (2013) had a larger sample size (model development n = 2240) and validation n = 2200), the dataset used was for women admitted to ICU and the EWS included the fraction of inspired oxygen and arterial blood gas, these may limit its use in a low resource setting ^25, 27, 28^. We did not have the fraction of inspired oxygen in our dataset but found that low SPO2 was not a significant predictor of SMO risk in our final model. It would have been useful to assess that performance on our dataset, but this was impossible because the clinical variables in the SMO cases of our dataset were collected as categorical (binary) variables.

We found that temperature >38° C was a predictor of SMO, similar to findings of Ryan et al. (2015)^29^ and Singh et al. (2012)^15^. Our finding of high systolic BP as a predictor of SMO was consistent with findings of two inpatient obstetric ward-based validation studies which used obstetric morbidity^15,^ as defined by consensus of experts, and ICU admission as outcomes^28^, and an EWS external validation study that had death as outcome ^19^. These findings differed from the development and validation study by Carle and colleagues (2013), that used data within 24 hours of admission into ICU, while the other studies used data from inpatients that had no obvious need for critical care ^18,26^.

Our report of a significant association between SMO and tachypnoea, high pulse rate, diastolic blood pressure, and low consciousness level was consistent with the findings from the ICU-based external validation study by Paternina-Caicedo et al.^19^.

We found a significantly increased risk of SMO among women who had caesarean compared to vaginal birth, and this remained an important predictor of SMO in all our models during development. This informed our inclusion of mode of birth in the proposed EWS tool (Table 5 and Additional file: Appendix 2).

Our models have excellent predictive ability to discriminate women who developed SMO from those who did not (AUCs consistently above 90%). The model attained similar diagnostic predictive accuracy as one developed, internally^18^ and externally^19^ validated using data from obstetric ICU patients in the USA. Our model also performed similarly to non-obstetric cardiovascular, adult critical care and neonatal critical care score systems. 29–31 Our models have significantly better screening characteristics (PPV 94% CI 93-95) as compared to an average of 41% reported for 16 different EWS versions. ^7^ This is of particular significance, since an EWS that generates many false-positive findings may worsen clinical care, constituting a nuisance alarm by creating an excessive burden on health systems^32,33^. Potentially, the proposed obstetric EWS algorithm presents an opportunity to institute life-saving interventions to improve clinical outcome. However, current evidence suggests that the use of EWS by itself is not enough to improve health outcomes and that for this tool to perform optimally, an EWS must be integrated with an outreach support system, such as a rapid response team ^7,34^.

Of the seven parameters in the proposed EWS algorithm (Table 5 and Additional file: Appendix 2), five (temperature, pulse rate, systolic blood pressure, respiratory rate and consciousness level) were included in the majority (>80%) of EWSs published to date^7^. Delays in triage (identification of who is, or may become, severely ill and should be provided with a higher level of care) are believed to contribute immensely to an increased burden of adverse obstetric outcomes in this settings^5^. This is further confounded by the unavailability of patient monitoring devices and other diagnostic equipment, especially in primary healthcare settings ^28^. Therefore, in addition to inpatient obstetric wards, we believe the proposed algorithm can present a potentially useful triaging tool to aid timely referral in primary healthcare centres in LMICs. However, prospective external validation is recommended to assess the effectiveness of this tool in primary, secondary and tertiary care in other low resource settings to substantiate these recommendations.

Main strengths of our analysis lie in the robust maternal deaths and near-misses data set which was prospectively collected primarily for research purposes with very few missing data (2·7% of participants), and strict adherence to a diagnostic predictive model development process and reporting as recommended by TRIPOD.^10^ A limitation of EWS validation studies that were addressed in our analysis is lack of standardisation of outcome measures; this is especially common with studies using morbidity as outcome measures, as often, this was defined based on consensus rather than standardised definitions^7^. There are several limitations to this study. Firstly, data of SMO consisted of already categorised clinical variables. Although cut-offs were based on recommendations for defining specific disease conditions (such as high blood pressure) by policy-making organisations like WHO, it was not possible to validate different trigger thresholds for the model parameters. Secondly, lack of continuous data also made it impossible to externally validate other EWS versions like CEMACH’s MEOWS and the ICU based chart developed by Carle et al. (2013)^18^. Additionally, pulse oximetry was poorly recorded in our study data set, especially in controls where the parameter was assumed to be above 90%. Although the absence of oxygen saturation could make our proposed chart more feasible to use in low resource settings, evidence from other analyses has shown that it is a valuable predictor of death and serious obstetric complications^35–38^. This clinical variable would therefore probably still contribute to a statistically developed and validated EWS decision tool in our study population. This can be investigated in an appropriately designed study in the future. Finally, the performance of our model might have been overestimated by the case-control study design with a high proportion of SMO cases. External validation should preferably be performed in a cohort study with a representative incidence of SMO (2.7% as in the Nigerian near-miss study).

## Conclusion

To the best of our knowledge, this study provides for the first time an internally validated statistically developed predictive model for SMO among all women admitted to obstetric wards in a low resource setting. This model was used to develop a simple score-based EWS algorithm that has easy to measure parameters with readily available patient monitoring tools, hence constituting a potentially useful triaging tool in low resource healthcare settings. Further work is, however, needed to validate this proposed chart externally in obstetric wards as well as primary healthcare settings.

## Data Availability

The datasets used during the current study are available from the corresponding author on reasonable request.

## List of abbreviations

AUC: Area under the curve
AUROC: Area under the receiver operator characteristic curve
AVPU: Alert, responds to voice or pain and unresponsive
CEMACH: United Kingdom Confidential Enquiry into Maternal and Child Health
DBP: Diastolic blood pressure
EWS: Early warning scores
ICU: Intensive care unit
LMIC: Low and middle-income countries Millenium Development Goals
MEOWS: Modified early obstetric warning scores
MMR: Maternal Mortality Ratio
ROC: Receiver operator characteristic
RR: Respiratory rate
SBP: Systolic blood pressure
SMO: Severe maternal outcomes
TRIPOD: Transparent Reporting of a multivariable prediction model for Individual Prognosis Or Diagnosis
WHO: World Health Organization

## Declaration

### Ethics approval and consent to participate

Clearance was obtained from the principal investigators of the Nigerian Near-Miss study (from the WHO, Geneva, Switzerland) for the use of the data. A data use agreement was signed in which the PIs indicated that this analysis was covered under the approved use of data by the Ethics Review Committee at the WHO and all 42 participating hospitals. However, additional approval for this study was obtained from the Research Ethics Committee of the Liverpool School of Tropical Medicine (Protocol ID 17-047).

## Consent for publication

Not applicable

## Availability of data and materials

The datasets used and/or analysed during the current study are available from the corresponding author on reasonable request.

## Authors’ contributions

All authors have read and approved the manuscript. AU: study design, analysis, interpretation, drafting and review. AM: statistical input for design, analysis, interpretation and review. MM: study design, interpretation and review, CA: conceptualisation, study design, interpretation, drafting and review.

## Acknowledgement

We acknowledge the profound support of Dr Olafemi T Oladapo and the Nigeria Near-miss and Maternal Death Surveillance Network for providing the SMO data set used in this analysis. We also thank Dr U. S. Bawa (Ahmadu Bello University Teaching Hospital, Zaria-Nigeria), Professor Calvin M. Chama (Abubakar Tafawa Balewa University Teaching Hospital, Bauchi-Nigeria) and Dr Idris Liman (National Hospital Abuja, Nigeria) for their support during our control data collection.

## Additional files

Appendix 3: Receiver-operating curve for the prediction of severe maternal outcome in the developmental model (n = 600)

Appendix 4: Receiver-operating curve for the prediction of severe maternal outcome in the validation model (n = 900)

Appendix 5: Receiver-operating curve for the prediction of severe maternal outcome in the sensitivity model with total dataset (n=5243)

